# An iterative supervised learning method identifying two subgroups of FOLFOX resistance patterns and predicting FOLFOX response in colorectal cancer patients

**DOI:** 10.1101/2020.06.10.20127167

**Authors:** Sun Tian, Fulong Wang, Shixun Lu, Rujia Wu, Gong Chen

## Abstract

**Background:** FOLFOX is a combination of drugs that is widely used to treat colorectal cancer. The response rate of FOLFOX in colorectal cancer(CRC) is 30-50%. We develop a method that analyzes mechanisms of FOLFOX resistance and predicts whether a patient will benefit from FOLFOX.

**Methods:** Gene expression data of 83 stage IV CRC tumor samples (FOLFOX responder n=42, non-responder n=41) were used to develop a supervised learning method IML and analyze subgroups of FOLFOX resistance mechanism. Datasets of 32 FOLFOX treated stage IV CRC patients and 55 FOLFOX treated stage III CRC patients were used as independent validations.

**Results:** An iterative supervised learning (IML) method identified two distinct subgroups of CRC patients who resist FOLFOX. Each subgroup relies on different types of DNA damage repair proteins and they are mutually exclusive. Protein-protein networks showed the main mechanism might be the synergistic effect of resisting apoptosis and an altered cell cycle. IML method was validated in two independent validation sets, one FOLFOX treated stage IV CRC patients(HR=2.6, p-value=0.02, 3-years survival rate of the predicted responder group 61.9%, predicted nonresponder group 18.8%) and one FOLFOX treated stage III CRC patients (estimated HR=2.36, p-value=0.02). A subgroup of mesenchymal subtype patients shows the pattern as FOLFOX responders.

**Conclusions:** IML method reflects the underlying biology of FOLFOX resistance and predicts FOLFOX response.

## Background

Despite the recent advance of cancer immunotherapy, chemotherapy remains to be the backbone of treatment for the majority of colorectal cancer patients. FOLFOX is a combination of chemotherapy drugs comprising leucovorin, 5-FU, and oxaliplatin, and FOLFOX is widely used to treat colorectal cancer.[1] The observed objective response to FOLFOX in metastatic colorectal cancer is approximately 30-50%.[2, 3] To select colorectal cancer patients that will likely benefit from or resist to FOLFOX treatment will allow choosing the most effective treatment at the beginning and avoid unnecessary side effects for patients. It is therefore important to develop methods to stratify FOLFOX responders and FOLFOX nonresponders. To predict FOLFOX response is not a trivial task, and the complexity is at three levels. Firstly, FOLFOX is not a single targeted therapy with known single protein targets. The main cytotoxic components of FOLFOX, 5-Fu and oxaliplatin, have their own different mechanism of action. 5-FU aims to inhibit thymidylate synthase that is a key enzyme in DNA synthesis, and the metabolites of 5-FU have similar structure as nucleotides can be incorporated into DNA and lead to cell death.[4] Oxaliplatin forms intra-strand links between DNA and disrupts DNA replication.[5] Secondly, colorectal cancer is a heterogeneous disease and at least four consensus molecule subgroups exist.[6] The dependence on DNA replication and cell cycle of colorectal tumor cells in different molecule subgroups are not the same. Thirdly, tumors likely resist FOLFOX treatment in different ways.[7] Thus the resistance is likely caused by a combination of different factors and the typical simple sequencing one drug target gene approach will unlikely work.[7]

Machine learning methods such random forest, support vector machine, and neural network developed using gene expression data have been proved useful in predicting FOLFOX treatment response.[3, 8] The general procedures of these machine learning approaches is to compare all FOLFOX nonresponders with all FOLFOX responders and identify most significantly differentially expressed signature genes between these two groups, then tune parameters of algorithms to reach the optimal performance. While these procedures are technically solid from machine learning perspective, they rely on one biological assumption: when one uses all FOLFOX nonresponders to compare with all FOLFOX responders to retrieve signature genes in the first place, the fundamentally biological assumption is that all FOLFOX nonresponders share the same mechanism of FOLFOX resistance. However, colorectal cancer is a heterogeneous disease and the assumption that all FOLFOX resistance tumors largely share a common character is unlikely true.

In this report, using genomic data of colorectal cancer patients with objective FOLFOX response data, we develop an iterative supervised learning (IML) method that identifies main subgroups of colorectal cancer patients who share the same mechanism of FOLFOX resistance. For each subgroup, we analyzed FOLFOX resistance mechanism separately and showed that each subgroup displays its own different unique underlying biology of FOLFOX resistance. Score functions were constructed for each subgroup and then combined. Scores of IML method can not only select patients who benefit from FOLFOX treatment, but also indicate the mechanism of resistance. We validated IML method in two independent validation datasets: one FOLFOX treated stage IV colorectal cancer patients and one FOLFOX treated stage III colorectal cancer patients.

## Results

### Development of IML model and two subgroups of FOLFOX resistance patterns using stage IV CRC tumors

The overall design of the machine learning flow is illustrated in figure 1 (**Figure 1**). The machine learning method resulted in statistical converged points in the third and the sixth iterative supervised learning rounds, with which the average area under curve values of 200 rounds of 10-folder cross-validation became consistently higher than 0.8. The third and sixth iterative supervised learning round were used to select genes in the final signature. In the third iterative supervised learning round, all 42 FOLFOX responder samples and a subset of 30 FOLFOX non-responder samples were used for the statistical analysis (methods section for detailed statistical screens), and a 74 gene signature *S*_1_ was trained and represents biological characters of the first subgroup of FOLFOX resistance pattern (**Gene signature 1, Table 1**). In the sixth iterative supervised learning round, all 42 FOLFOX responder samples and a subset of 13 FOLFOX non-responder samples were used for the statistical analysis, and a 74 gene signature *S*_2_ was trained and represents biological characters of the second subgroup of FOLFOX resistance pattern (**Gene signature 2, Table 1**).

**Figure 1.**
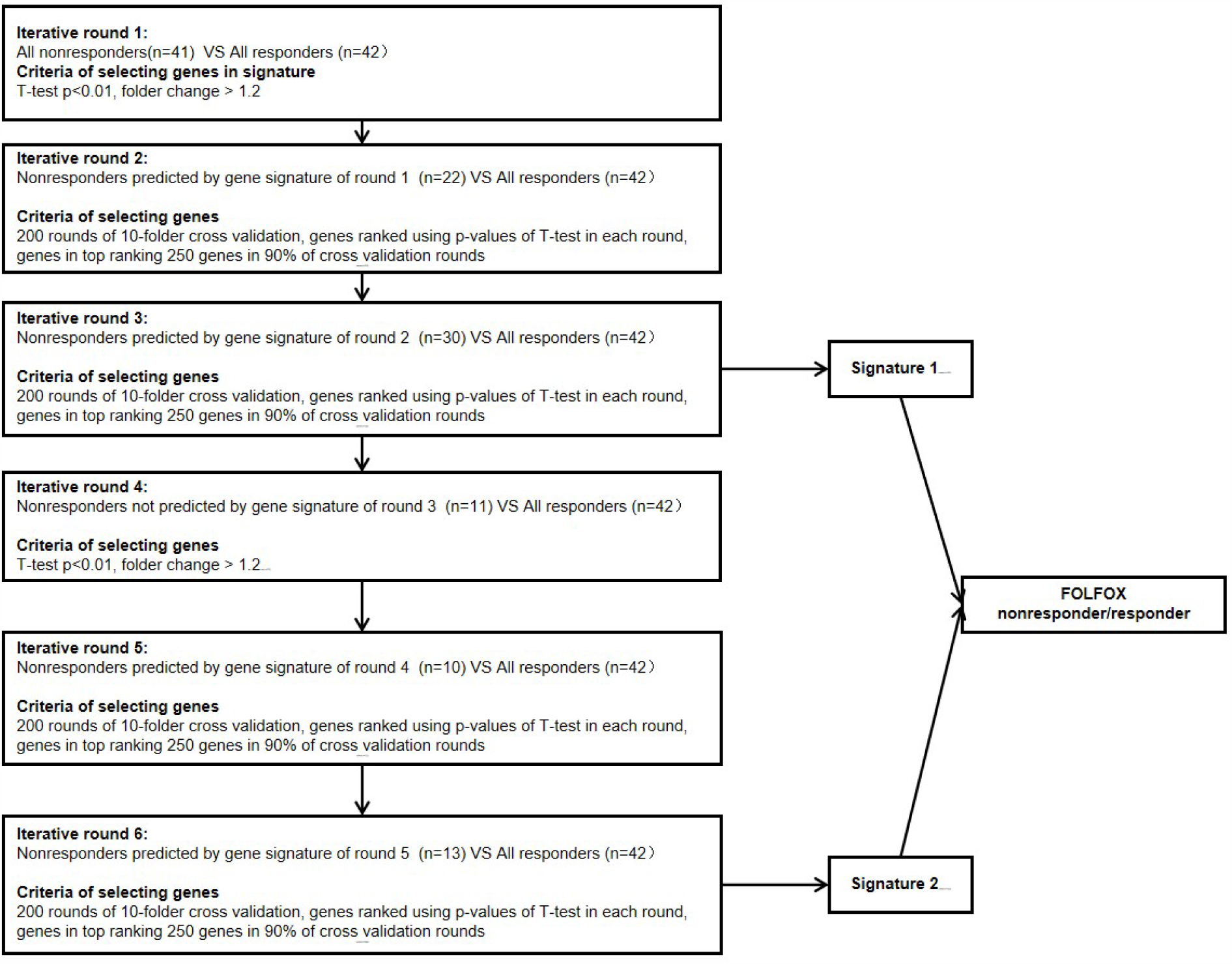
The workflow of iterative supervised learning method finds convergence points of the machine learning process, divided FOLFOX nonresponders to two subgroups, and then analyze the FOLFOX resistance mechanism of two subgroups of FOLFOX nonresponders.

**Table 1A.**
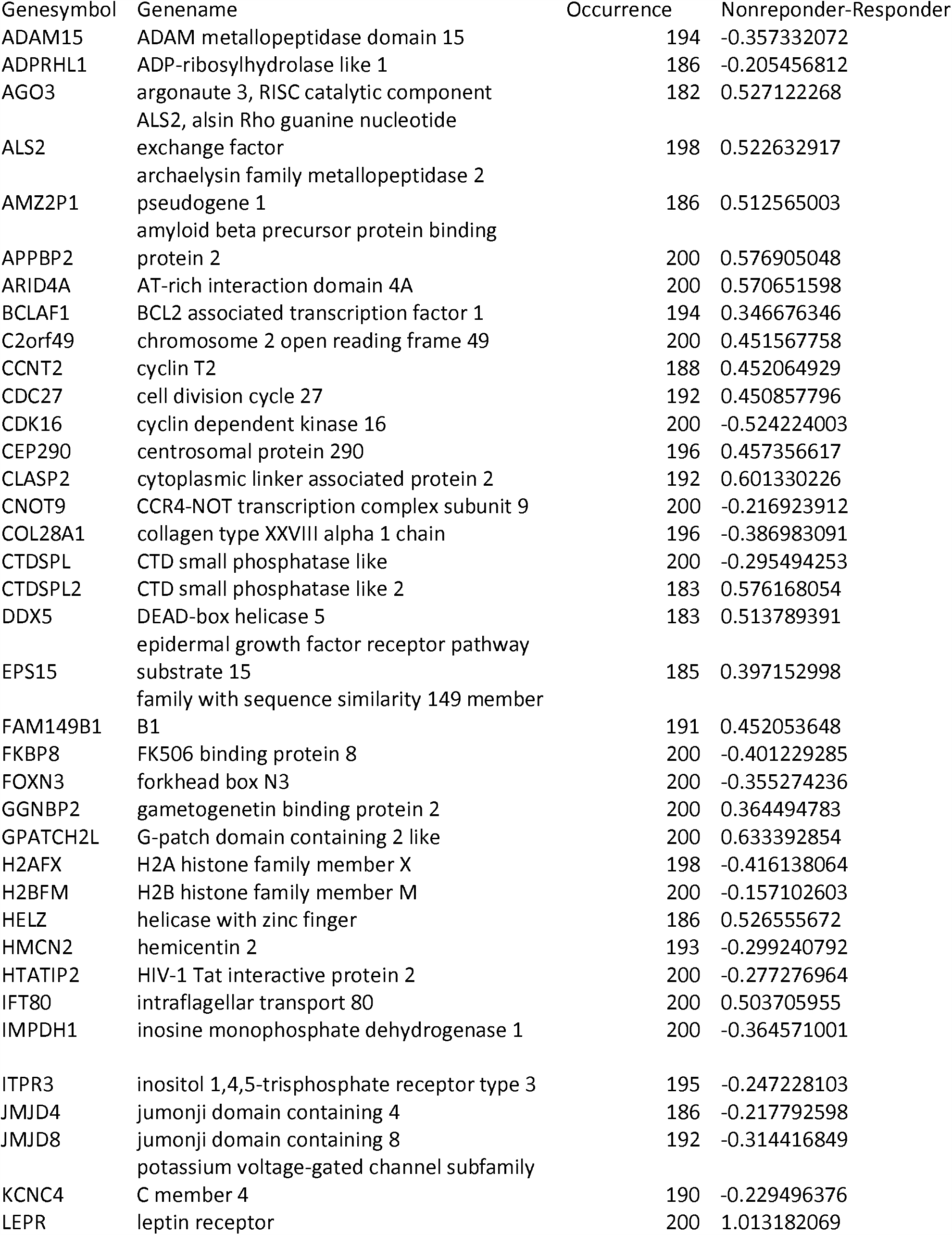
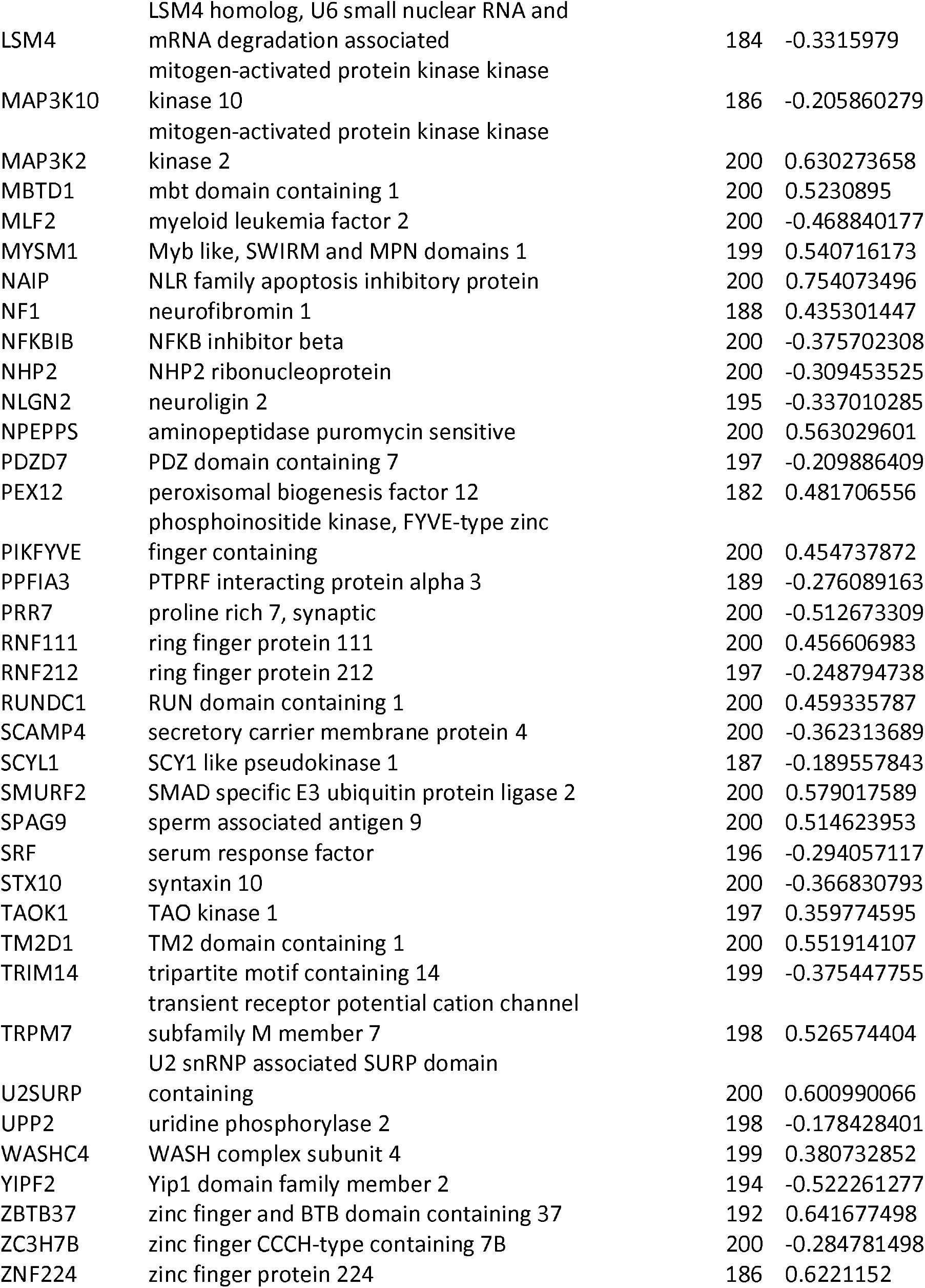
List of genes in IML signature 1

| Genesymbol | Genename | Occurrence | Nonreponder-Responder |
| --- | --- | --- | --- |
| ADAM15 | ADAM metallopeptidase domain 15 | 194 | -0.357332072 |
| ADPRHL1 | ADP-ribosylhydrolase like 1 | 186 | -0.205456812 |
| AGO3 | argonaute 3, RISC catalytic component | 182 | 0.527122268 |
| ALS2 | ALS2, alsin Rho guanine nucleotide exchange factor | 198 | 0.522632917 |
| AMZ2P1 | archaelysin family metallopeptidase 2 pseudogene 1 | 186 | 0.512565003 |
| APPBP2 | amyloid beta precursor protein binding protein 2 | 200 | 0.576905048 |
| ARID4A | AT-rich interaction domain 4A | 200 | 0.570651598 |
| BCLAF1 | BCL2 associated transcription factor 1 | 194 | 0.346676346 |
| C2orf49 | chromosome 2 open reading frame 49 | 200 | 0.451567758 |
| CCNT2 | cyclin T2 | 188 | 0.452064929 |
| CDC27 | cell division cycle 27 | 192 | 0.450857796 |
| CDK16 | cyclin dependent kinase 16 | 200 | -0.524224003 |
| CEP290 | centrosomal protein 290 | 196 | 0.457356617 |
| CLASP2 | cytoplasmic linker associated protein 2 | 192 | 0.601330226 |
| CNOT9 | CCR4-NOT transcription complex subunit 9 | 200 | -0.216923912 |
| COL28A1 | collagen type XXVIII alpha 1 chain | 196 | -0.386983091 |
| CTDSPL | CTD small phosphatase like | 200 | -0.295494253 |
| CTDSPL2 | CTD small phosphatase like 2 | 183 | 0.576168054 |
| DDX5 | DEAD-box helicase 5 | 183 | 0.513789391 |
| EPS15 | epidermal growth factor receptor pathway substrate 15 | 185 | 0.397152998 |
| FAM149B1 | family with sequence similarity 149 member B1 | 191 | 0.452053648 |
| FKBP8 | FK506 binding protein 8 | 200 | -0.401229285 |
| FOXN3 | forkhead box N3 | 200 | -0.355274236 |
| GGNBP2 | gametogenetin binding protein 2 | 200 | 0.364494783 |
| GPATCH2L | G-patch domain containing 2 like | 200 | 0.633392854 |
| H2AFX | H2A histone family member X | 198 | -0.416138064 |
| H2BFM | H2B histone family member M | 200 | -0.157102603 |
| HELZ | helicase with zinc finger | 186 | 0.526555672 |
| HMCN2 | hemicentin 2 | 193 | -0.299240792 |
| HTATIP2 | HIV-1 Tat interactive protein 2 | 200 | -0.277276964 |
| IFT80 | intraflagellar transport 80 | 200 | 0.503705955 |
| IMPDH1 | inosine monophosphate dehydrogenase 1 | 200 | -0.364571001 |
| ITPR3 | inositol 1,4,5-trisphosphate receptor type 3 | 195 | -0.247228103 |
| JMJD4 | jumonji domain containing 4 | 186 | -0.217792598 |
| JMJD8 | jumonji domain containing 8 | 192 | -0.314416849 |
| KCNC4 | potassium voltage-gated channel subfamily C member 4 | 190 | -0.229496376 |
| LEPR | leptin receptor | 200 | 1.013182069 |
| LSM4 | LSM4 homolog, U6 small nuclear RNA and mRNA degradation associated | 184 | -0.3315979 |
| MAP3K10 | mitogen-activated protein kinase kinase kinase 10 | 186 | -0.205860279 |
| MAP3K2 | mitogen-activated protein kinase kinase kinase 2 | 200 | 0.630273658 |
| MBTD1 | mbt domain containing 1 | 200 | 0.5230895 |
| MLF2 | myeloid leukemia factor 2 | 200 | -0.468840177 |
| MYSM1 | Myb like, SWIRM and MPN domains 1 | 199 | 0.540716173 |
| NAIP | NLR family apoptosis inhibitory protein | 200 | 0.754073496 |
| NF1 | neurofibromin 1 | 188 | 0.435301447 |
| NFKBIB | NFKB inhibitor beta | 200 | -0.375702308 |
| NHP2 | NHP2 ribonucleoprotein | 200 | -0.309453525 |
| NLGN2 | neuroligin 2 | 195 | -0.337010285 |
| NPEPPS | aminopeptidase puromycin sensitive | 200 | 0.563029601 |
| PDZD7 | PDZ domain containing 7 | 197 | -0.209886409 |
| PEX12 | peroxisomal biogenesis factor 12 | 182 | 0.481706556 |
| PIKFYVE | phosphoinositide kinase, FYVE-type zinc finger containing | 200 | 0.454737872 |
| PPFIA3 | PTPRF interacting protein alpha 3 | 189 | -0.276089163 |
| PRR7 | proline rich 7, synaptic | 200 | -0.512673309 |
| RNF111 | ring finger protein 111 | 200 | 0.456606983 |
| RNF212 | ring finger protein 212 | 197 | -0.248794738 |
| RUNDC1 | RUN domain containing 1 | 200 | 0.459335787 |
| SCAMP4 | secretory carrier membrane protein 4 | 200 | -0.362313689 |
| SCYL1 | SCY1 like pseudokinase 1 | 187 | -0.189557843 |
| SMURF2 | SMAD specific E3 ubiquitin protein ligase 2 | 200 | 0.579017589 |
| SPAG9 | sperm associated antigen 9 | 200 | 0.514623953 |
| SRF | serum response factor | 196 | -0.294057117 |
| STX10 | syntaxin 10 | 200 | -0.366830793 |
| TAOK1 | TAO kinase 1 | 197 | 0.359774595 |
| TM2D1 | TM2 domain containing 1 | 200 | 0.551914107 |
| TRIM14 | tripartite motif containing 14 | 199 | -0.375447755 |
| TRPM7 | transient receptor potential cation channel subfamily M member 7 | 198 | 0.526574404 |
| U2SURP | U2 snRNP associated SURP domain containing | 200 | 0.600990066 |
| UPP2 | uridine phosphorylase 2 | 198 | -0.178428401 |
| WASHC4 | WASH complex subunit 4 | 199 | 0.380732852 |
| YIPF2 | Yip1 domain family member 2 | 194 | -0.522261277 |
| ZBTB37 | zinc finger and BTB domain containing 37 | 192 | 0.641677498 |
| ZC3H7B | zinc finger CCCH-type containing 7B | 200 | -0.284781498 |
| ZNF224 | zinc finger protein 224 | 186 | 0.6221152 |

**Table 1B.**
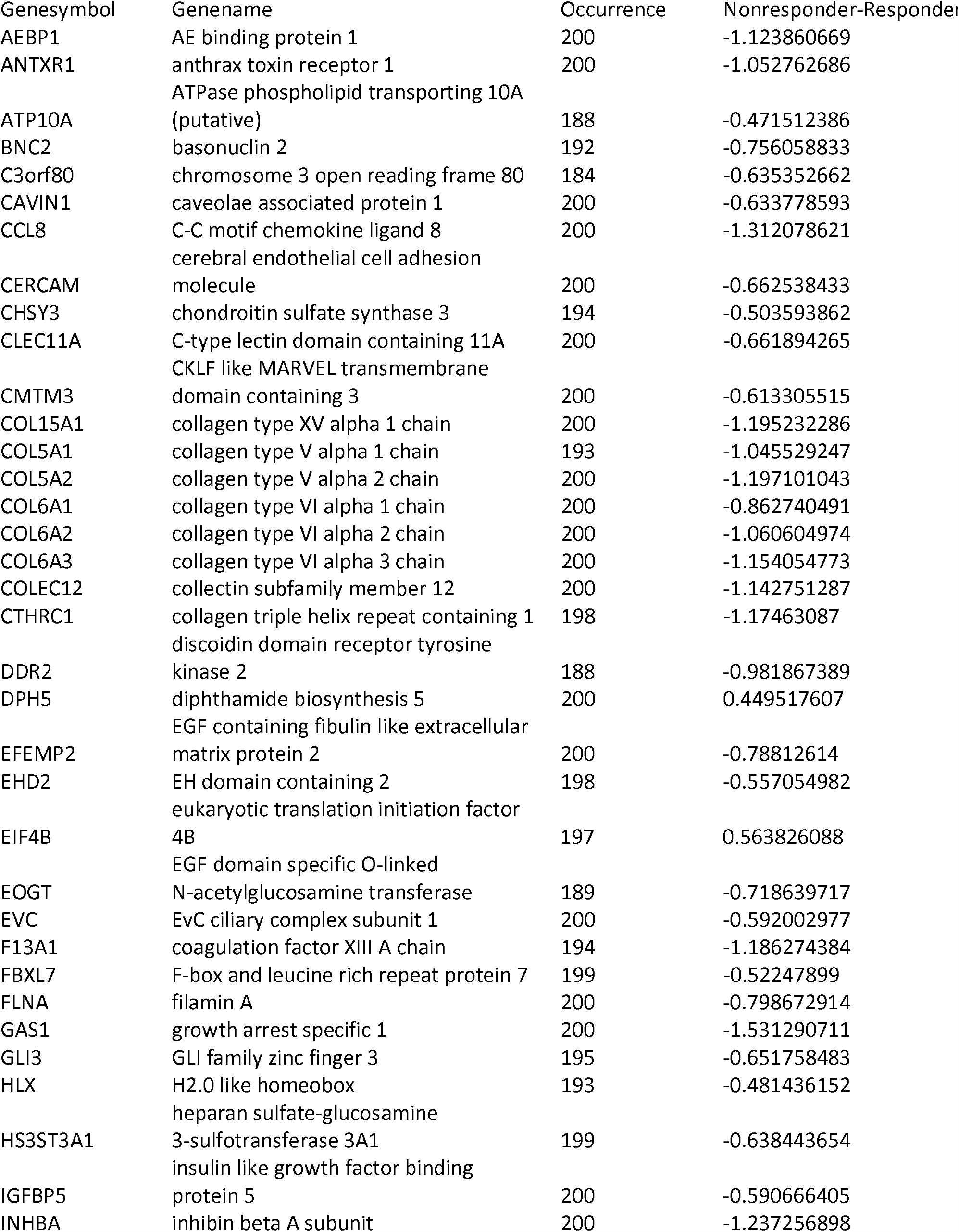
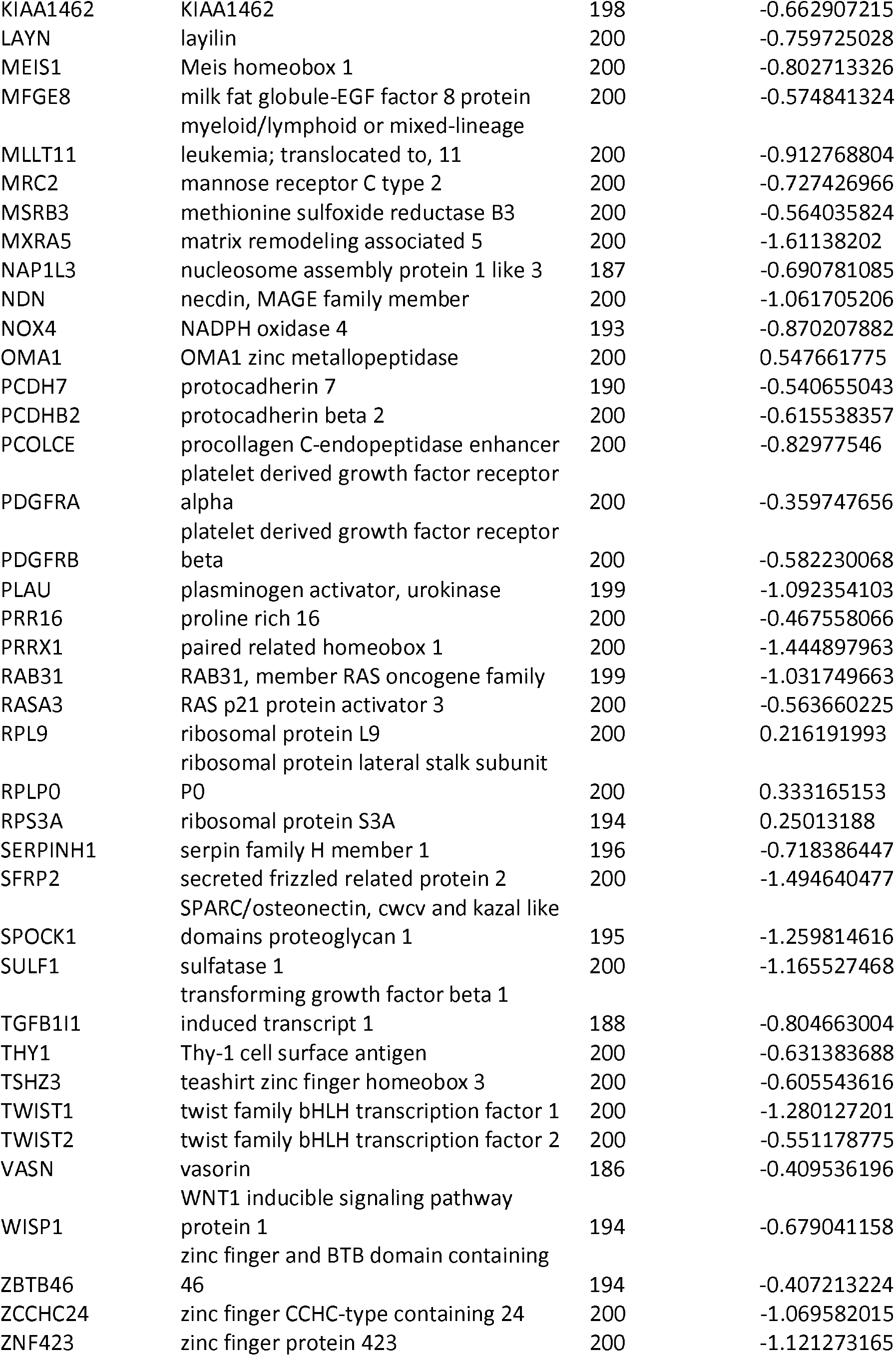
List of genes in IML signature 2

### Performance of IML model in the training set and independent validation set

In the training set of 83 FOLFOX treated stage IV CRC tumor samples (GSE28702, responder n=42, non-responder n=41), the IML prediction model resulted in a sensitivity of 97.6% and a specificity of 100%. The overall survival data of the training set is not available, thus the survival curve was not calculated for the training set.

In the independent validation set of 32 FOLFOX treated stage IV CRC samples (GSE72970), overall survival data is available. 50% of the samples (n=16) were predicted by the IML model as FOLFOX non-responder and 50% of the samples (n=16) were predicted by the IML model as FOLFOX responders. Despite the small sample size, the survival analysis of the IML prediction model resulted in a significant hazard ratio HR=2.6 (p-value=0.02). The predicted FOLFOX responder group has a 3-years survival rate 61.9% [95% CI, 41.9%-91.4%] and the predicted FOLFOX non-responder group has a 3-years survival rate 18.8% [95% CI, 6.8%-52.0%]. The inferred 95% confidence interval is large is due to the small sample size (n=32). The median overall survival time of the predicted FOLFOX non-responder group (13.4 months) is significantly shorter than the predicted FOLFOX responder group (36.6 months) (**Figure 2**).

**Figure 2.**
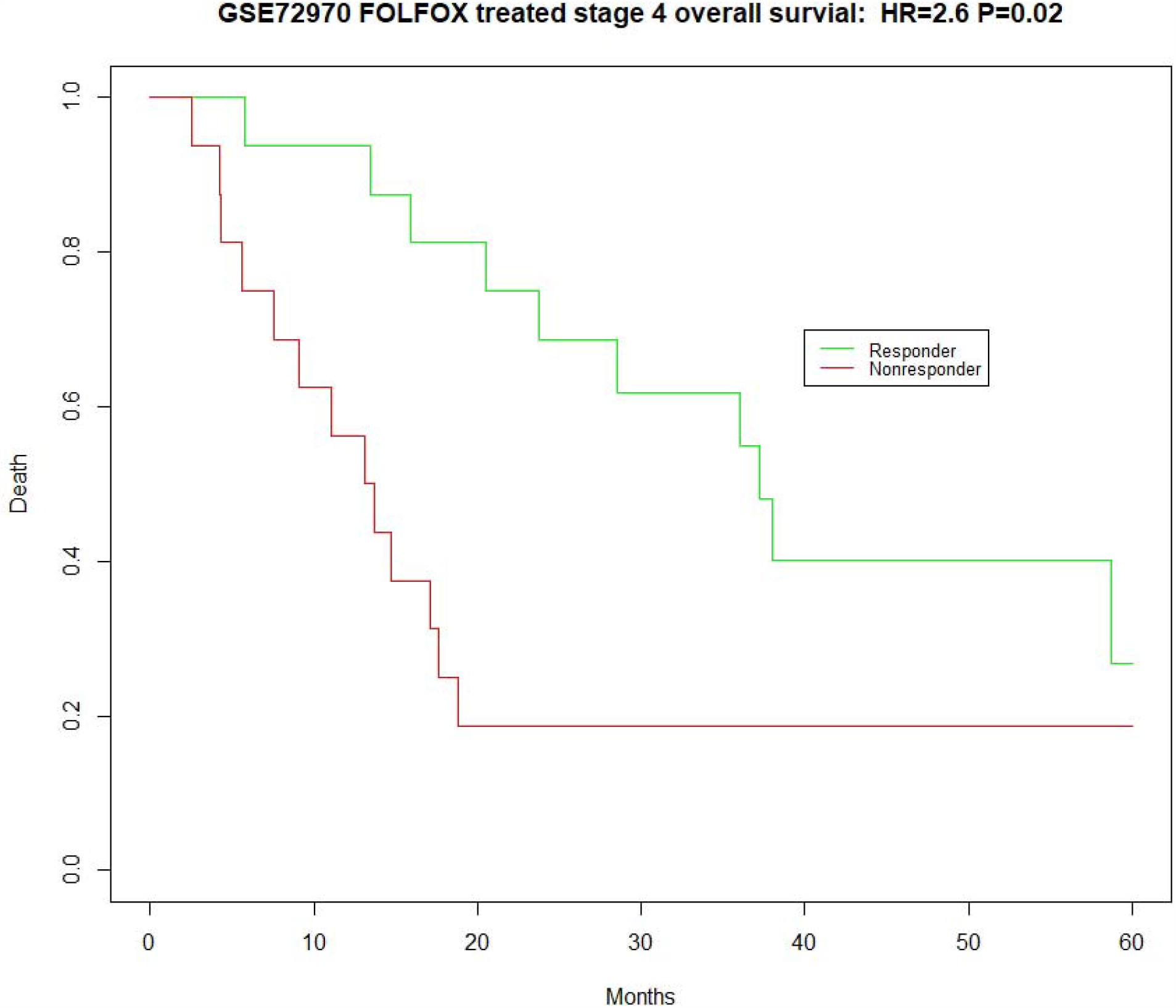
Survival curve of FOLFOX treated stage 4 colorectal cancer patients (n=32) in the validation set GSE72970 showed the IML predicted FOLFOX responder group has significantly better overall survival than the IML predicted FOLFOX nonresponder group HR=2.6 (p-value=0.02)

To test the predictive power of the IML prediction model in the adjuvant setting, we used a cohort of 166 stage III colorectal cancer patients who received FOLFOX as adjuvant therapy (GSE81653). The complexity to use data of FOLFOX treated stage III colorectal patient samples to test the predictive power of a treatment response model developed using stage IV patient samples is to separate the co-founding prognostic factors from the real treatment response factor. To remove the potential bias introduced by intrinsic prognostic characters of tumors, we used CMScaller to classified 166 stage III colorectal cancer patients into four CMS consensus molecular subgroups (CMS1 n=16, CMS2 n=32, CMS3 n=19, CMS4 n=55, Unclassified n=44).[6, 9] Because the CMS4 group tends to have the worst prognosis, and within this cohort of stage III patients, the CMS4 group also has the largest number of patients (n=55) for statistical analysis, and we used only consensus molecular subgroups CMS4 subgroups of patients to test the response to FOLFOX treatment. The recurrence events of patients in this dataset are available in the GEO database. The follow-up time of patients of this dataset is not available and is estimated as an evenly distributed time series over 60 months. The IML model showed good predictive power in these 55 FOLFOX treated stage III patients, the hazard ratio of predicted non-responder group to responder group is estimated as HR=2.36 (p-value=0.02) (**Figure 3**). Despite the common belief that CMS4 mesenchymal tumors tend to resist chemotherapy, it is important to note that the IML method can still identify subgroups of patients from CMS4 subgroups that benefit from FOLFOX treatment. The IML method is specifically developed for the prediction of FOLFOX response, and it provides additional predictive value.

**Figure 3.**
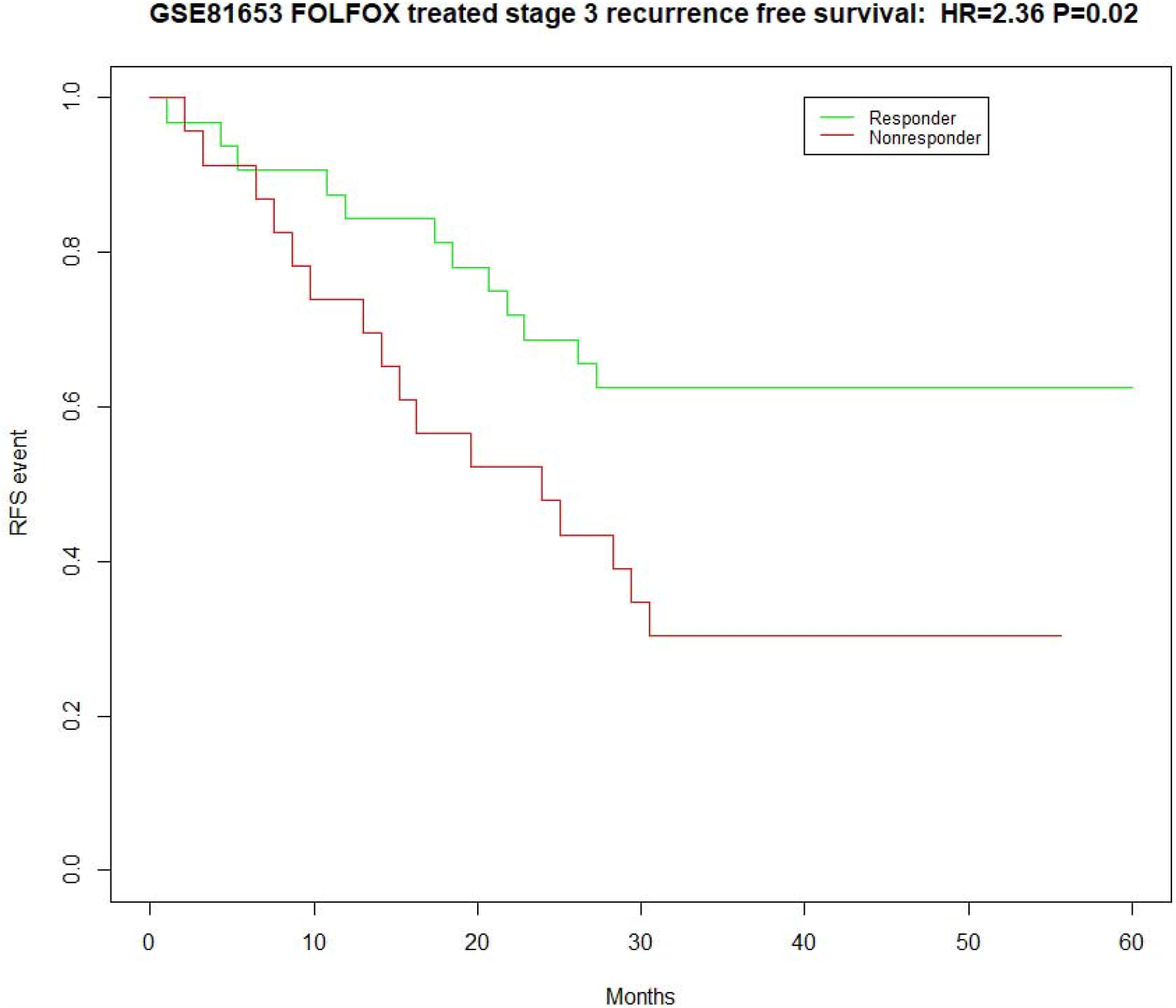
In the adjuvant setting, the survival curve of FOLFOX treated stage 3 colorectal cancer patients (n=55) in the validation set GSE81653 showed the IML predicted FOLFOX responder group showed significantly better recurrence free survival than the IML predicted FOLFOX nonresponder group HR=2.36 (p-value=0.02). The recurrence events of patients in this dataset were recorded. The follow-up time of patients of this dataset was not available and was estimated as an evenly distributed time series over 60 months.

### Molecular mechanism and protein-protein interaction network of two subgroups of FOLFOX resistance

The underlying FOLFOX resistance mechanisms of two different resistance subgroups are different. The cluster of GO biological process term of enriched function analysis of genes in signature *S*_1_ indicates apoptotic process (Enrichment score 0.805, **Enriched function of gene signature 1, Table 2A**). The scores of FOLFOX nonresponders predicted by signature 1 showed a tendency of high ERCC1 and high DPYD, suggesting tumor cells in major percentages of FOLFOX nonresponders might have relative high catabolism rate of 5-FU and have an efficient nucleotide excision repair by ERCC1 to overcome to the apoptosis induced by oxaliplatin (**red box, Figure 4A, Figure 4B**). Further, the protein interaction network analysis of genes in signature *S*_1_ showed a highly interconnected network of cell cycle related proteins (**Figure 5**) and enriched GO terms of proteins participate interaction are strongly cell cycle and mitosis associated functional terms (**Table 3**). Taken together, these results indicated that the dominant mechanism of FOLFOX resistance might be the synergistic effect of the intrinsic ability of tumor cells themselves to resist apoptosis and have an altered cell cycle.

**Figure 4.**
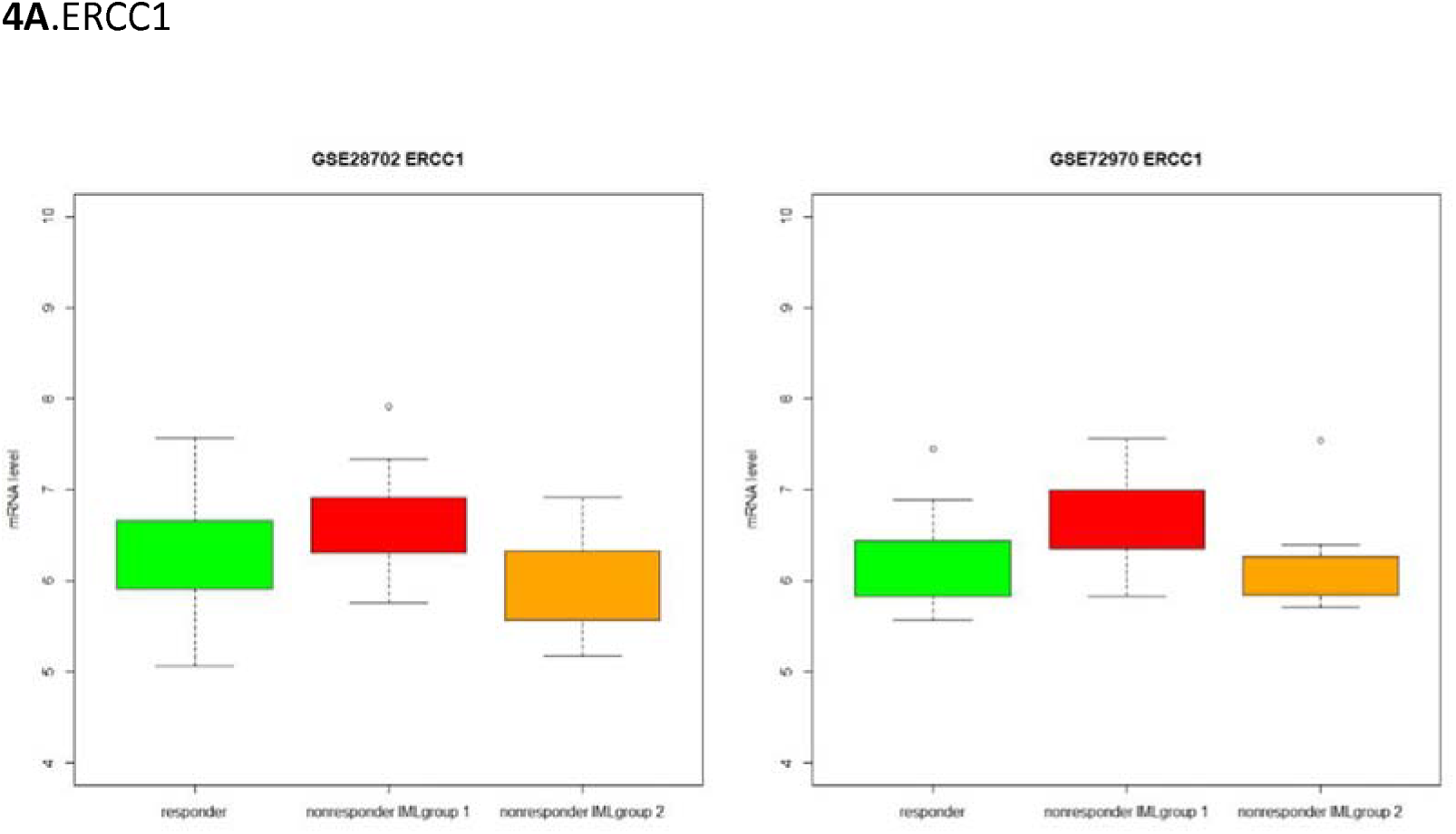
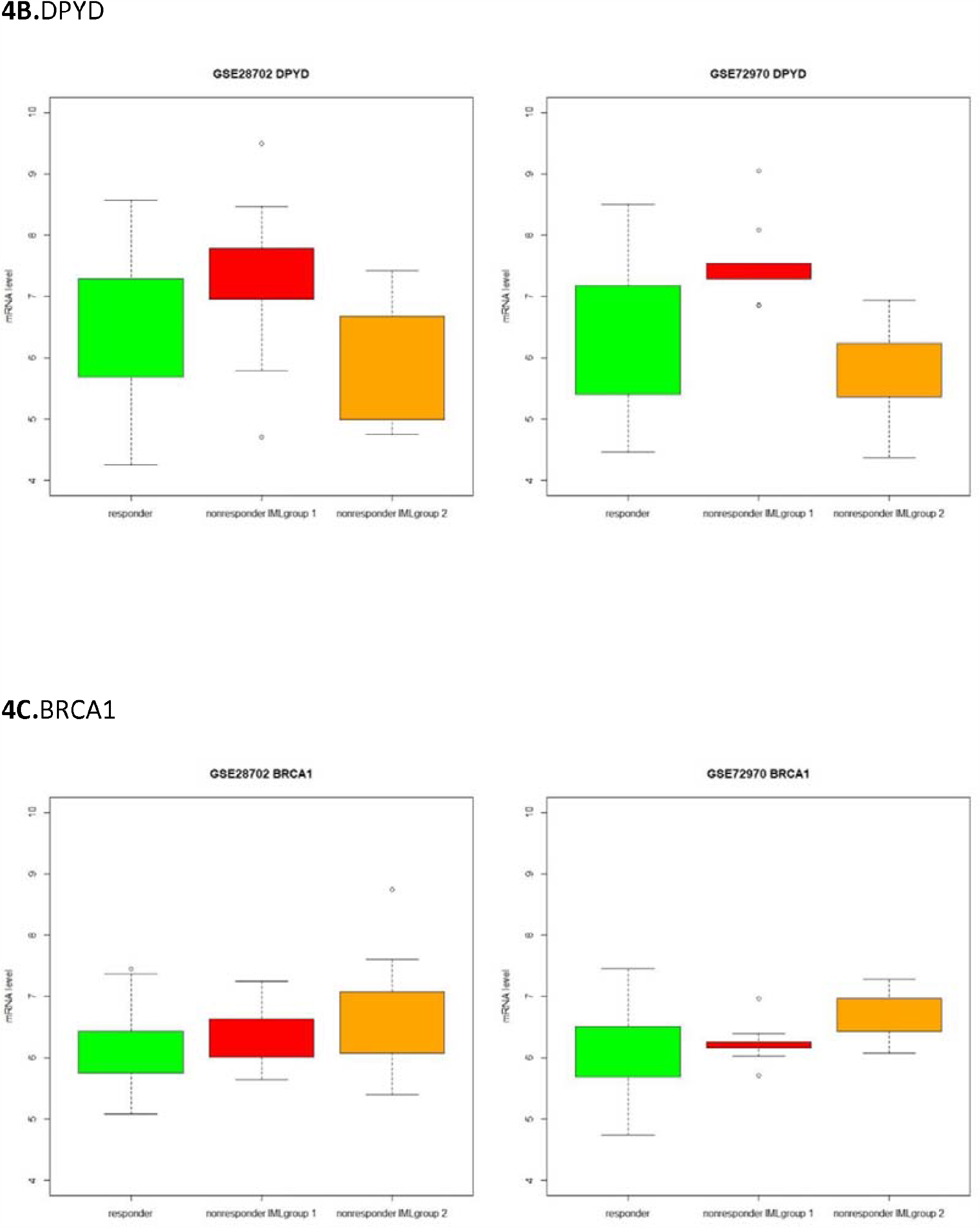
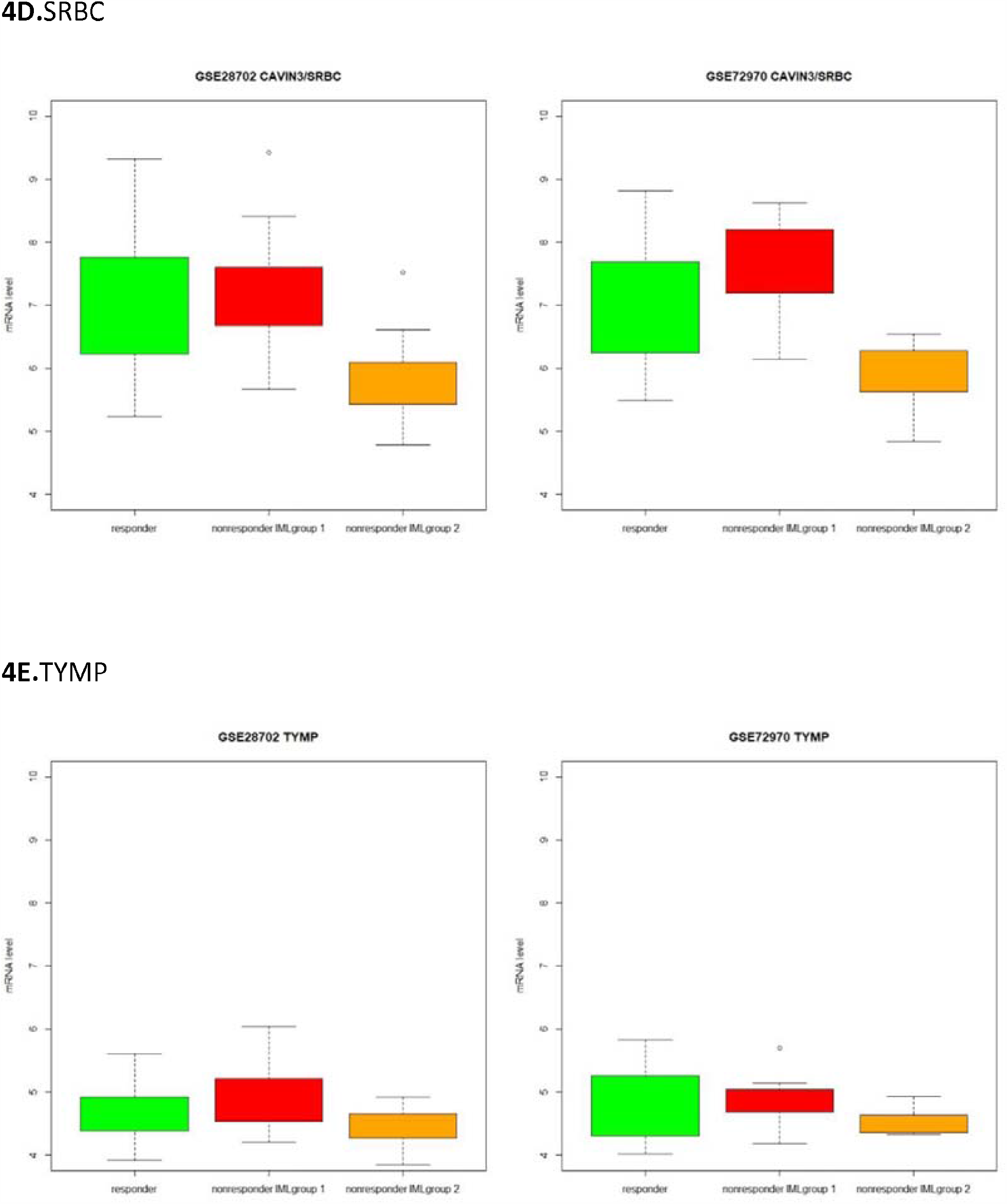
Boxplots showed associations between expression levels of known markers (**4A.**ERCC1, **4B.**DPYD, **4C.**BRCA1, **4D.**SRBC, **4E.**TYMP) of FOLFOX resistance with the IML prediction results. Y-axis is expression values of this marker, green boxes are IML predicted FOLFOX responders, red boxes are IML signature 1 predicted FOLFOX nonresponders, and yellow boxes are IML signature 2 predicted FOLFOX nonresponders. The underlying major FOLFOX resistance mechanisms of two subgroups are different.

**Figure 5.**
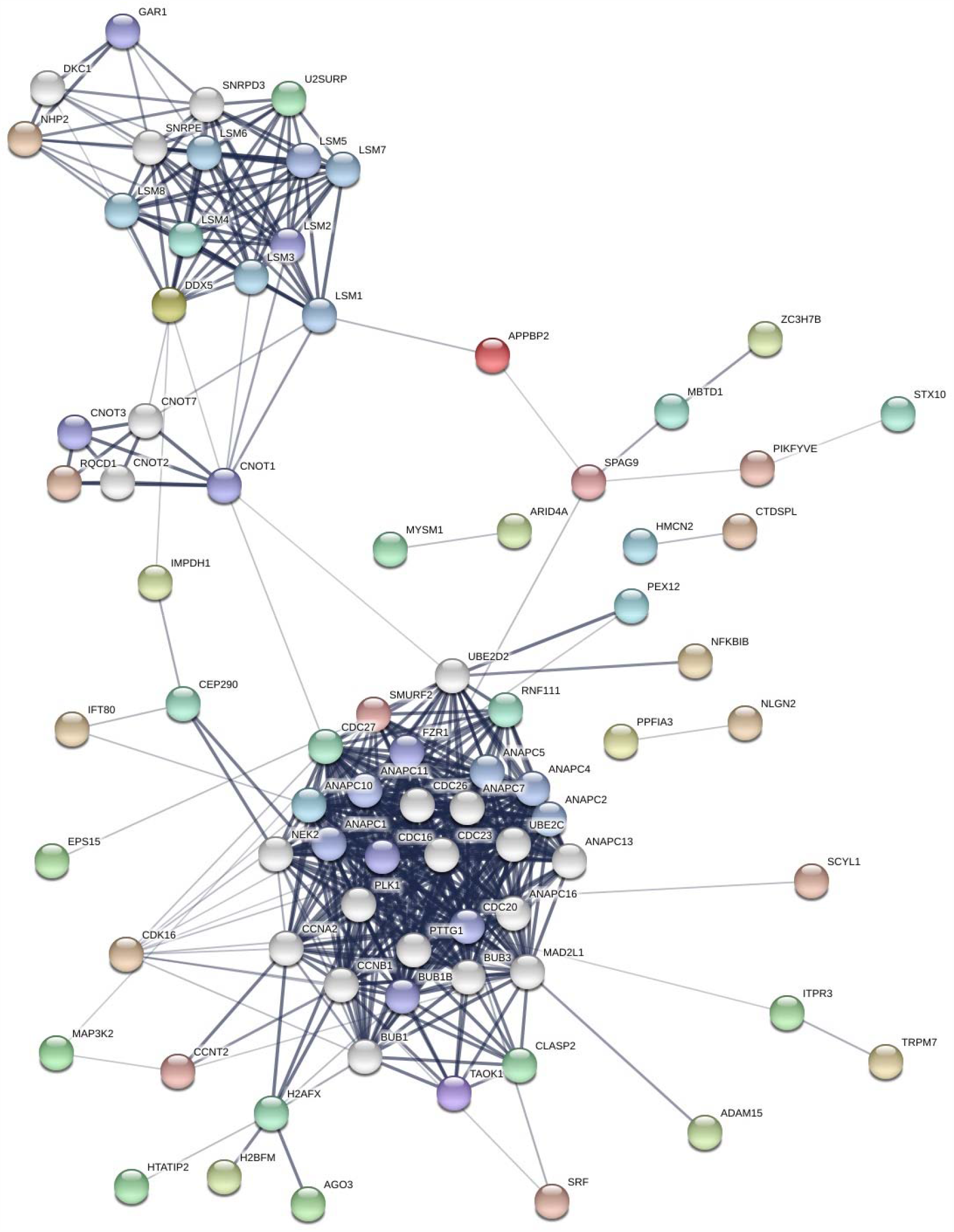
Protein interaction network of genes in the IML signature 1 showed a highly connected cell cycle and mitosis related network.

**Table 2A.** Clusters of enriched functional GO terms of genes in IML signature 1 This table is too long to put in the main manuscript file, please see it in the supplementary files

**Table 2B.** Clusters of enriched functional GO terms of genes in IML signature 2 This table is too long to put in the main manuscript file, please see it in the supplementary files

**Table 3.** Enriched functional GO biological process terms (pvalue < 0.01) of all genes participating protein-protein interaction network of IML signature 1. This table is too long to put in the main manuscript file, please see it in the supplementary files

The resistant mechanism suggested by enriched function analysis of genes in the signature *S*_2_ is less clear (**Enriched function of gene signature 2, Table 2B**). Overall, the downregulation of genes such as SPOCK1, TGFB1I1, WISP1, TWIST1, TWIST2 in nonresponders in the signature *S*_2_ displayed a clear suppression of the mesenchymal phenotype. The activation of mesenchymal phenotype was reported to resist chemotherapy.[6, 10] Here, the suppression of the mesenchymal phenotype is observed positively associated with the resistance to FOLFOX. This observation does not mean that suppression of the mesenchymal phenotype is the cause of resistance to FOLFOX, rather, the observed suppression of the mesenchymal phenotype is likely to be the effect of functional DNA damage repair proteins. The scores of FOFOX nonresponders predicted by signature *S*_2_ showed an association of high BRCA1 and low level of BRCA1 inactivator CAVIN3, suggesting tumor cells in signature *S*_2_ subgroup of FOLFOX nonresponders may rely on BRCA1 to repair double strand breaks induced by oxaliplatin (**yellow box, Figure 4C, Figure 4D**). The pattern of activated BRCA1 is consistent with the absence of a mesenchymal phenotype as BRCA1 is known to suppress epithelial to mesenchymal transition and stem cell dedifferentiation.[11] In addition, the FOLFOX nonresponders predicted by signature *S*_2_ showed a mild tendency of low TYMP (**yellow box, Figure 4E**), suggesting tumor cells in signature *S*_2_ subgroup of FOLFOX nonresponders might have a less efficient conversion from 5-FU to its active metabolite 5-FdUMP.

## Discussion

In this report, we present the IML model that identifies two subgroups of colorectal cancer patients who have distinct mechanisms of FOLFOX resistance. IML model can predict response to FOLFOX treatment and this model was validated in both FOLFOX treated stage IV patients and FOLFOX treated stage III patients. The main advantage of the IML model is the underlying design assumes there can be multiple causes of FOLFOX resistance in colorectal cancer patients. Our results suggest that there are at least two different major mechanisms of FOLFOX resistance in colorectal cancer. These two mechanisms depend on the upregulation of different types of DNA damage repair proteins and they are largely mutually exclusive. The dominant mechanism of FOLFOX resistance is the synergistic effect of anti-apoptosis and altered cell cycle of tumor cells, and represents approximately 75% of the nonresponders. The second mechanism of FOLFOX resistance is featured by the activation of BRCA1, and represents approximately 25% of the nonresponders. These two mechanisms of FOLFOX resistance are largely mutually exclusive.

The signatures of the IML model showed correlations with known single gene markers such as ERCC1, DPYD, BRCA1, CAVIN3, and TYMP. However, it should be noted that a single gene cannot fully explain FOLFOX resistance, and resistance is caused by the combination of various factors.[7] These observed correlations between signatures with known single gene markers are consistent in samples of both training set GSE28702 and validation set GSE72970 (**Figure 4A, Figure 4B, Figure 4C, Figure 4D, Figure 4E**), indicating that the underlying biology of resistance identified by the IML method is robust, and the IML model indeed captures the gene expression pattern of the synergistic effect of resistance to apoptosis, altered cell cycle, dysfunction of drug metabolism and unregulated DNA repair that contribute to FOLFOX resistance.

Epithelial-mesenchymal subtype is thought to tend to resist chemotherapy. However, within 55 stage III patients of epithelial-mesenchymal subtype, the IML method identified a subgroup of patients with epithelial-mesenchymal subtype who could still benefit from FOLFOX treatment. These results not only suggest that the IML method provides specific additional predictive value for FOLFOX response, but also indicates that epithelial-mesenchymal transition alone might not be sufficient to cause resistance to FOLFOX treatment in all tumors. The acquisition of synergistic effects of anti-apoptosis and the upregulation of different types of DNA damage repair proteins are also required and this is likely to be an independent process of epithelial-mesenchymal transition.

The limitation of our study is the sample size of FOLFOX treated CRC tumors in the publicly available database. The total sample size used in this study is 170. Although this number is indeed moderate, the underly biological pattern retrieved by IML model is shown to be robust, and performance in the independent validation sets is statistically significant. This demonstrated one concept that a predictive biomarker needs to be designed based on the understanding of underlying biology, rather than merely relying on large samples, statistical powers and computational power to optimize parameters of advanced machine learning algorithms. A predictive biomarker dissecting underlying biology of drug response, even using a relatively small sample size, still could find its root in biology and show robust results.

## Materials and Methods

### Data

R/Bioconductor software was used to analyze gene expression data.[12] Publicly available gene expression data and objective FOLFOX response data of 83 stage IV colorectal cancer patients (GSE28702) were used as the training set.[3] Gene expression data and survival data of 32 FOLFOX treated stage IV colorectal cancer patients (GSE72970) were used as the independent validation set in the metastatic setting.[13] Gene expression data and survival data of 55 FOLFOX treated stage III colorectal cancer patients (GSE81653) were used as the independent validation set in the adjuvant setting.[14] Normalization of Affymetrix Human Genome U133 Plus 2.0 Array data (GSE28702 and GSE72970) was performed using the frozen RMA (fRMA) method in *frma* package, and this normalization method is designed for the clinical diagnostic settings that each single samples is processed individually.[15] The batch effects of samples in GSE28702 and samples in GSE72970 were removed using *ComBat*.[16] The normalized Affymetrix Human Gene 2.0 ST Array data (GSE81653) was downloaded from Gene Expression Omnibus database. Patients or the public were not involved in the design, or conduct, or reporting, or dissemination plans of our research.

### Iterative supervised learning (IML) method

In total, 83 stage IV CRC tumor samples in dataset GSE28702 (FOLFOX responder n=42, FOLFOX non-responder n=41) were used to train the model. As showed in figure 1 (**Figure 1**), the model was trained using six iterative rounds. In each round, a selected subset of samples in the non-responder group was compared with all tumor samples in the responder group. The first and fourth learning rounds were used to preselect non-responder tumors with the same character in a subset, and a t-test of the non-responder group and the responder group used in this round were performed. Two statistical criteria were used to select genes in signatures of these two learning rounds: (1) genes with a p-value <0.01 and (2) the absolute value of the difference between the mean of the non-responder group and the responder group of genes needs to be higher than 1.2 folder change. The second, third, fifth and sixth learning round were used to reinforce the previous learning rounds, and 200 rounds of 10-fold cross-validation were performed. In each cross-validation round, a t-test was performed and p-values of genes were ranked. One statistical criterion was used to select genes in signatures of these four learning rounds: p-values of a gene were in the top 250 ranking genes in at least 90% of 200 rounds of cross-validations. The gene signatures selected in the first, second, fourth and five rounds were pre-screen signatures and they were not used in the final score functions. The scores of these four pre-screen signatures were calculated by using a simple nearest centroid model. The gene signatures selected in the third round and sixth rounds were final signatures and constructed two k-nearest neighbors regression score functions *S*_1_ and *S*_2_. The score function *S*_1_ and *S*_2_ reflect different biological characters of subgroups of FOLFOX resistance pattern. The two scores were combined to a single score *S* by combinatory rule *S* = (***S***_1_ ≥ *λ*_1_)ϒ(***S***_2_ ≥ *λ*_2_). A CRC tumor sample can be output into two groups: FOLFOX responder (*S* = 0) and FOLFOX non-responder (*S* = 1). Enriched function analysis of signatures was performed using DAVID.[17] Protein network analysis was performed using STRING.[18]

## Conclusion

We have developed the IML method that can stratify the FOLFOX responders and nonresponders. The IML method was validated in both stage III and stage IV colorectal cancer patient groups. The major advantage of our approach is that IML does not treat all FOLFOX resistant patients the same, rather, it unbiasedly stratifies the subgroups of nonresponders that share the same mechanism of the synergistic effect of FOLFOX resistance. The prediction score of the IML method reflects underlying molecular mechanisms of FOLFOX resistance. Different ways of FOLFOX resistance by tumors may need to be combatted by using FOLFOX in combination with other drugs.

## Data Availability

Data generated for the current study are available from the corresponding author on reasonable request.

## Authors’ contributions

Design and concept: ST and GC. Sample collection and data quality: FW, SL. Project planning: RW. Data analysis and statistics: ST and RW. Write the first draft: ST. Read and review the final paper: ST, FW, SL, RW and GC.

## Ethics approval and consent to participate

No human or animal ethics approval was required for this study.

